# Assessment of Knowledge for Urinary Tract Infections Among Pregnant Women in Jordan: A Cross-Sectional Study

**DOI:** 10.64898/2026.03.06.26347768

**Authors:** Sara H. Alawdat, Zeinab M. Hassan

## Abstract

**Background & Aims:** Urinary tract infections (UTIs) are common health issue during pregnancy, often lead to adverse maternal and neonatal outcomes if left untreated, low knowledge contribute to high UTI rates, particularly in resource-limited settings like Jordan. The aims it’s to assess the knowledge levels about UTIs among pregnant women in Jordan and its association with socio-demographic characteristics.

**Methods:** A descriptive cross-sectional study was conducted among 500 pregnant women attending antenatal clinics in four major governmental hospitals across Jordan. Data were collected using a validated questionnaire based on the Theory of Planned Behavior (TPB) comprising 25 questions, including 5 socio-demographic questions and 20 knowledge questions, scores were categorized as “adequate” or “inadequate” based on the median score, associations between categorical variables, such as socio-demographic factors and levels of knowledge, were examined using chi-square tests. For continuous variables, t-tests and ANOVA were used to compare means across groups, such as comparing knowledge scores across different educational levels or income groups.

**Results:** Among participants, 51.4% had inadequate knowledge, while 48.6% demonstrated adequate knowledge. Higher knowledge levels were significantly associated with younger age (21–30 years), urban residence, higher education (university and postgraduate), and employment status.

**Conclusion:** The findings highlight a knowledge gap among pregnant women regarding UTIs. Integrating targeted health education and addressing socio-demographic disparities into antenatal care, especially for women with low education and rural residence, may improve maternal outcomes.

## Introduction

Pregnancy is a transformative period in a woman’s life, filled with excitement and anticipation, but also demands heightened attention to health and well-being to ensure positive outcomes for both the mother and fetus [1]. Urinary tract infections (UTIs) are the most prevalent infections during pregnancy, affecting up to 23.9% of pregnant women globally [2]. Urinary tract infections (UTIs) happen when bacteria, often from the skin or rectum, enter the urethra and infect the urinary tract, with bladder infections (cystitis) being the most prevalent [1]. The hormonal and mechanical changes during pregnancy, such as increased progesterone levels and uterine pressure on the bladder, create an environment conducive to bacterial growth and urinary stasis, further increasing the risk of infection [3]. The primary causative agent of UTIs is Escherichia coli, accounting for 56.45% of cases among pregnant women, followed by Klebsiella pneumonia (14.31%) and Proteus mirabilis (7.62%) [4]. These infections are associated with a range of complications, including preterm birth, low birth weight, and preeclampsia, which can have lasting effects on maternal and neonatal health [5,6]. The most common symptoms include nocturia, urgency of micturition, bladder pain, and nocturnal enuresis, all of which can significantly affect the quality of life during pregnancy [7].

Knowledge plays a critical role in enabling pregnant women to recognize symptoms, adopt preventive practices, and seek early treatment [8]. Despite the high prevalence of UTIs in low- and middle-income countries, few studies have focused on the knowledge level among pregnant women in Jordan.

This is study was designed to answer the following research questions:

- What is the level of knowledge of UTI among pregnant women?
- Is there an association between the knowledge level of UTI and socio-demographic variables among pregnant women?

## Methods

### Study design and setting

This was a descriptive cross-sectional study conducted from October 2023 to July 2024 in four major governmental hospitals in Jordan: Al-Bashir Hospital in Amman, Al-Karak Governmental Hospital in Karak, Princess Badea Governmental Hospital in Irbid, and Al-Zarqa Governmental Hospital in Zarqa. These facilities were conveniently chosen to ensure a representative sample from across the country.

### Study population and sample

The target population for this study consisted of pregnant women attending antenatal and gynecology clinics, total of 500 pregnant women were recruited using convenience sampling. These clinics operate daily, offering antenatal care to pregnant women, including nutrition and dietary counseling, as well as the provision of recommended supplements such as folic acid and iron. The sampling method used for this study was convenience sampling, a commonly employed technique in clinical and population research. Inclusion criteria included being currently pregnant, aged between 18–50 years, and attending antenatal clinics at the selected hospitals, women with chronic renal conditions or congenital urinary anomalies were excluded.

### Instrument

The questionnaire was developed by researcher and was created and utilized in Iran in 2013, it was based on the constructions of the theory of planned behavior (TPB) [9]. The questionnaire consisted of two sections: socio-demographic included five information of respondents (pregnant women); age, educational level, occupation, residence, family income. The second deals with knowledge of pregnant women about UTI and prevention with (20 questions) designed as yes, no, and (There is no answer); the correct option received a score of 2, and the wrong option received a score of 0 for no, and (There is no answer). The knowledge scores ranged from 0 to 40, based on the median; the knowledge level was divided into two levels: inadequate knowledge, below the median, and adequate knowledge, above the median. The language of the questionnaire was Persian, and it was translated into English and then into Arabic by bilingual experts according to the steps of the World Health Organization [10]. Modifications were made, corrections were conducted accordingly to suit the Jordanian culture, and tools were designed in their final format. The tool validity was assessed by four-panel experts in maternity nursing, adult nursing, and obstetrics medicine, who reviewed the tools for clarity, relevance, and comprehensibility, the total CVI for the tool was 0.82 which is considered highly valid. A pilot study was done on 10% of the estimated sample size by the researcher, the translated questionnaires were distributed among 50 pregnant women in antenatal clinics at Al-Bashir Hospital; these participants were excluded from the study population Some modifications were required to be made to all sections of the questionnaire. The internal consistency of the questionnaire was evaluated using the Statistical Package for Social Sciences (SPSS) by (Cronbach’s alpha) test. The results were: Cronbach’s alpha for reproductive history = 0.84, Cronbach’s alpha for knowledge = 0.964. Internal consistency is considered acceptable when the reliability coefficient is greater than or equal to.70 [11].

### Data collection procedures

Data were collected from October 2023 to July 2024, before data collection, the researcher gave an introduction and explained the study’s objectives to pregnant women who agreed to participate; participants were meted after they had received their antenatal care, and informed consent was obtained from each pregnant woman prior to her contribution to the study, only those who will be consented were recruited into the study and given a questionnaire, all data were collected in a comfortable, secluded room or in clinics at the hospital.

### Ethical considerations

Ethical approvals were obtained from the institutional review board (IRB) committees at Hashemite University (No. 10:2300508), and approval from the Ministry of Health (No. 13505) to conduct the research in the previously mentioned hospitals. Also, ethical permission to use the questionnaire was obtained. A consent form was introduced at the beginning of the questionnaire for the study. Acceptance of the participant was considered as consent; no rewards were provided to the respondents, and they had the right to withdraw without penalty at any moment as the participation was voluntary, the structured questionnaire was pre-tested and used as the data collection instrument, privacy was guaranteed despite the nature of the questionnaire.

### Data analysis

SPSS version 25 was used for data analysis. Descriptive statistics summarized demographic variables and knowledge levels, associations between categorical variables, such as socio-demographic factors and levels of knowledge, were examined using chi-square tests. For continuous variables, t-tests and ANOVA were used to compare means across groups, such as comparing knowledge scores across different educational levels or income groups.

## Results

### Knowledge Levels

Table 1 illustrates the distribution of participants according to their level of knowledge about urinary tract infections, out of 500 participants revealed that 51.4% (n=257) of pregnant women had inadequate knowledge, scoring below the median of 33 out of 40. In contrast, 48.6% (n=243) demonstrated adequate knowledge, with scores equal to or above the median, as illustrated in Figure 1 this highlights that slightly more than half of the participants lacked sufficient knowledge regarding UTIs.

**Table 1:** Knowledge level score about UTI (n=500).

| UTI Knowledge Levels | N | Percent |
| --- | --- | --- |
| Adequate Knowledge (≥ Median 33) | 243 | 48.6% |
| Inadequate Knowledge ($<$ Median33) | 257 | 51.4% |

**Figure 1.**
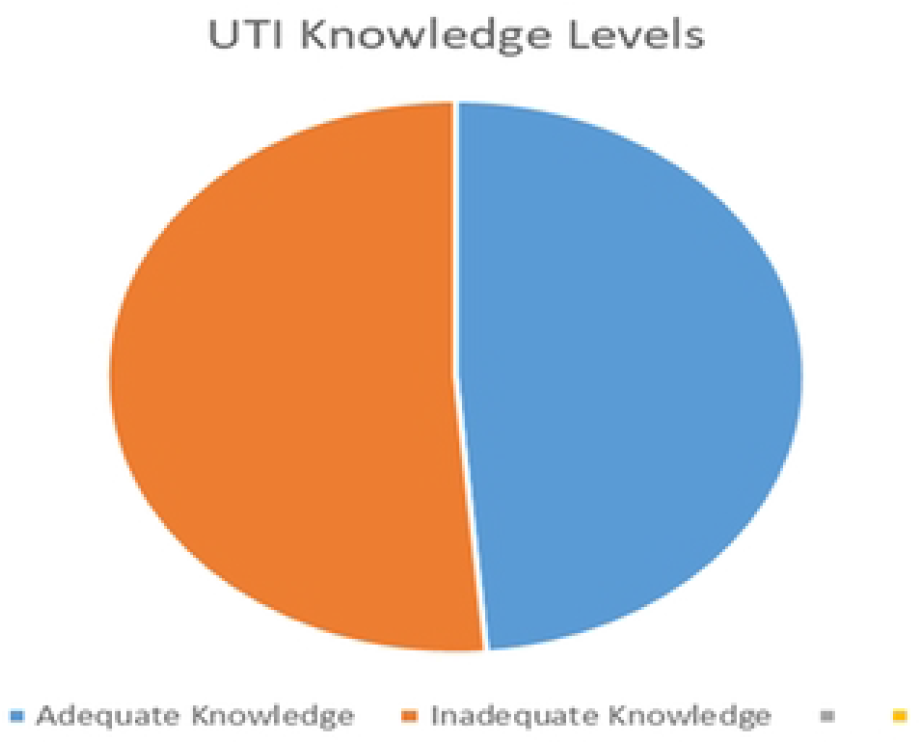
This figure displays that 51.4%, 48.6% of studied pregnant women had Inadequate and adequate knowledge respectively.

### Associations Between Level Knowledge of UTI and Socio- demographic characteristics

As shown in Table 2, the analysis of associations between knowledge levels about urinary tract infections (UTIs) and socio-demographic characteristics of the participants revealed several statistically significant relationships.

- Age (p < 0.01): Younger women (21–30 years) demonstrated higher knowledge levels compared to older women.
- Education level (p < 0.001): Participants with university or postgraduate education had significantly higher knowledge.
- Residency (p < 0.001): Urban residents showed better knowledge compared to rural residents.
- Employment (p = 0.02): Employed women had higher knowledge scores than unemployed women.
- Income level (p < 0.001): Participants with higher income had the greatest proportion of adequate knowledge, indicating that income plays a significant role in influencing UTI knowledge

**Table 2:** Associations Between Level Knowledge of UTI and Socio- demographic characteristics (n=500). (**P < 0.01) highly statistical significant difference.

| Socio- demographic characteristics | Adequate Knowledge | Inadequate Knowledge | p-value |
| --- | --- | --- | --- |
|  | N | N |  |
| Age |  |  |  |
| 18-20 Years | 9 | 30 | .001** |
| 21-30 Years | 129 | 112 |  |
| 31-40 Years | 93 | 89 |  |
| 41-50 Years | 12 | 26 |  |
| Educational level |  |  |  |
| Illiterate | 3 | 37 | .000** |
| Basic | 29 | 118 |  |
| Secondary | 35 | 81 |  |
| University | 165 | 21 |  |
| Postgraduate studies | 11 | 0 |  |
| Occupation |  |  |  |
| Working | 117 | 32 | .000** |
| Not working | 116 | 225 |  |
| Retired |  |  |  |
| Residence |  |  |  |
| Rural | 45 | 155 | .000** |
| Urban | 198 | 102 |  |
| Income |  |  |  |
| 260-460 JDs | 169 | 241 | .000** |
| 461-660 JDs | 72 | 13 |  |
| 661-860 JDs | 2 | 3 |  |
| More than 861 JDs | 0 | 0 |  |

## Discussion

The finding that 51.4% of participants had inadequate knowledge about urinary tract infections (UTIs) is a concerning indicator of gaps in health education among pregnant women in Jordan. UTIs are among the most common infections in pregnancy, yet inadequate understanding of their causes, prevention, and complications can lead to missed opportunities for early intervention and prevention. This inadequate knowledge underscores the need to reassess how information on UTIs is delivered during antenatal visits and through community health programs.

One potential justification for this finding is the variability in the quality and content of antenatal education provided in healthcare settings. In Jordan, although antenatal care is widely available, studies have pointed to inconsistencies in the health education component of these services [12,13]. Antenatal sessions often focus on broader topics such as fetal development and nutrition, with less emphasis on specific health risks like UTIs. Similarly, studies in Egypt demonstrated inadequate knowledge about UTIs due to the limited integration of infection prevention education in routine maternal care [14,15]. This reflects a pattern across many low- and middle-income countries (LMICs), where overburdened healthcare systems and resource constraints limit the depth and comprehensiveness of health education.

Similar results were found in the Philippines, where 77.39% of pregnant women had unsatisfactory knowledge about UTIs, citing cultural discomfort in discussing personal health issues [16]. These cultural barriers may prevent women from fully engaging in antenatal education sessions, even when such opportunities are available.

A comparative analysis with studies from other regions further contextualizes these findings. For instance, research in Pakistan found that 44.5% of pregnant women had insufficient knowledge about UTIs, while 55.5% demonstrated satisfactory knowledge [17]. This aligns closely with the results in Jordan but contrasts with findings in Nigeria, where reported that 64% of women had good knowledge about UTIs, likely due to targeted community health campaigns and better integration of UTI education into antenatal care [18]. Similarly, a study in Egypt found that 61.3% of Egyptian women had average knowledge, highlighting regional variations even within LMICs [14]. These differences suggest that targeted interventions, cultural adaptability, and the strength of public health systems significantly influence knowledge levels.

It is important to note that inadequate knowledge is not solely a reflection of the women’s capacity to learn but also of the healthcare system’s ability to effectively deliver information. Studies demonstrated that targeted educational interventions could significantly improve women’s knowledge about UTIs, in their quasi-experimental study, the proportion of women with good knowledge increased from 10% to 79.4% after an educational intervention [15,19]. This aligns with study which reported that self-care practices improved substantially when health literacy was addressed through structured educational programs [8].

Summing up, the finding that 51.4% of participants in this study had inadequate knowledge about UTIs underscores a critical need for targeted interventions in Jordan. Addressing this gap requires a multifaceted approach that includes improving the content and delivery of antenatal education, overcoming cultural barriers to communication, and addressing healthcare inequities that disproportionately affect rural and less educated populations. Comparative data from other regions highlight the importance of context-specific strategies, such as community outreach, health literacy programs, and standardized antenatal education protocols, to ensure that all women, regardless of their background, have access to the knowledge necessary to protect their health and that of their unborn children.

## Conclusion

In conclusion, this study sheds light on the critical issue of urinary tract infections (UTIs) among pregnant women in Jordan, revealing significant gaps in knowledge. The findings underscore the role of socio-demographic factors, such as age, education, occupation, residence, income, and chronic diseases, in influencing both knowledge. Addressing barriers such as resource limitations, misinformation, and disparities in healthcare access, particularly in rural and low-income settings, is essential. This study emphasizes the need for integrated interventions, enhanced antenatal education, and community-based strategies to bridge the knowledge-practice gap and reduce the prevalence and complications of UTIs among pregnant women, ultimately contributing to better maternal and neonatal health.

## Data Availability

All relevant data are within the manuscript and its Supporting Information files. Additional de-identified data are available from the corresponding author upon reasonable request and with approval from the Institutional Ethics Committee.

## Declarations

### Ethics Approval

Ethical approval was obtained from the Institutional Review Board of the Hashemite University and the Ministry of Health.

### Consent to Participate

Written informed consent was obtained from all participants.

### Consent for Publication

Not applicable; the manuscript does not contain any individual person’s data in any form.

### Availability of Data and Materials

The datasets generated and analyzed during the current study are available from the corresponding author on reasonable request.

### Authors’ Contributions

Sara H. Alawdat and Zeinab M. Hassan contributed substantially to the conception and design of the study. Sara H. Alawdat carried out the data collection, drafted the manuscript. Sara H. Alawdat and Zeinab M. Hassan analyzed and interpreted the data, reviewed the manuscript critically for important intellectual content. All authors read and approved the final manuscript and agreed to be accountable for all aspects of the work in ensuring that questions related to the accuracy or integrity of any part of the work are appropriately investigated and resolved.

## Acknowledgments

The author thanks all the pregnant women who participated in this study and the staff at the participating hospitals.

